# Increased whole body fluid volume status quantified by photon-counting detector CT in patients undergoing TAVR

**DOI:** 10.64898/2026.05.13.26352144

**Authors:** Nóra M Kerkovits, Miklós Vértes, Sámuel Beke, Scott Quadrelli, Péter Csákai-Szőke, A. Michael Peters, Lili Száraz, Ákos Varga-Szemes, Tilman Emrich, Bálint Szilveszter, Béla Merkely, Pál Maurovich-Horvat, Martin Ugander

**Affiliations:** Department of Radiology, Medical Imaging Centre, Semmelweis University, Budapest, Hungary; Kolling Institute, Royal North Shore Hospital, and University of Sydney, Sydney, Australia; Department of Nuclear Medicine, King’s College Hospital NHS Foundation Trust, London, United Kingdom; Department of Radiology and Radiological Science, Medical University of South Carolina, Charleston, SC, USA; Department of Diagnostic and Interventional Radiology, University Medical Center of the Johannes Gutenberg-University, Mainz, Germany; Heart and Vascular Center, Semmelweis University, Budapest, Hungary; Department of Clinical Physiology, Karolinska University Hospital, and Karolinska Institutet, Stockholm, Sweden

**Author notes:** **Corresponding author:** Martin Ugander, MD, PhD, Professor of Cardiac Imaging, Kolling Building, Level 12, Royal North Shore Hospital, St Leonards, Sydney NSW 2065, Australia. **Funding:** This study was funded in part from institutional research grant by Siemens Healthineers and from grants to Martin Ugander from the University of Sydney and Heart Research Australia. **Data sharing statement:** Data generated or analyzed during the study are available from the corresponding author by request.

## Abstract

**Background:** Before transcatheter aortic valve replacement (TAVR), patients with severe aortic valve stenosis are at an increased risk of developing fluid volume overload and heart failure, which is associated with subsequent adverse outcomes after TAVR.

**Purpose:** To quantify fluid volume status as whole-body fast-exchange extracellular volume (FE-ECV) in patients undergoing TAVR compared to healthy reference values using photon-counting CT (PCCT).

**Methods:** Consecutive patients referred for TAVR and healthy living kidney donor candidates, respectively, underwent PCCT including the pelvis. FE-ECV (mL) was quantified using venous hematocrit, injected iodinated contrast concentration and volume, and blood iodine concentration and urinary bladder excreted iodine mass quantified in iodine map regions of interest from the inferior vena cava and covering the urinary bladder, acquired at one time point 6-10 minutes after intravenous iodinated contrast administration.

**Results:** The study included 156 subjects (healthy: n=51, age 47±9 years, 55% female; TAVR: n=105, age 78±6 years, 39% female). In healthy subjects, FE-ECV was 160±22 mL/kg lean body mass (LBM), 95% limits 116–204 mL/kg LBM, and was independent of age, sex, contrast agent type, and scan delay time after contrast injection (p>0.66 for all). Compared to healthy subjects, FE-ECV in patients referred for TAVR was higher (174±34 mL/kg LBM, p=0.01), with 19 patients (18%) exceeding the normal range.

**Conclusion:** One in five patients referred for TAVR demonstrated increased FE-ECV, revealing a substantial prevalence of fluid overload detectable by single-time point late-phase PCCT iodine mapping.

**Summary statement:** PCCT iodine mapping is a simple method enabling objective quantification of whole-body fast-exchange extracellular volume, and revealing increased fluid volume status in patients with severe aortic stenosis undergoing TAVR.

**Key Results:**

1. Whole-body fast-exchange extracellular volume can be quantified using late-phase photon-counting CT iodine maps of the abdomen and pelvis.
2. Normal whole-body fast-exchange extracellular volume values in healthy subjects were determined for the method.
3. Patients with severe aortic stenosis scheduled for TAVR demonstrated increased whole-body fast-exchange extracellular volume.

## Introduction

In severe aortic stenosis, fluid overload can be assessed using bioimpedance, and reflects cardiac decompensation and is associated with an increased risk of heart failure and mortality^1^. Among patients referred for transcatheter aortic valve replacement (TAVR), excess fluid burden is linked to worse clinical outcomes^1,2^, and quantitative assessment of fluid overload using bioimpedance prior to intervention was shown to improve prediction of adverse post-interventional events beyond traditional measures of risk stratification^3^. Since excess fluid in this setting is predominantly retained in the extracellular space^4^, accurate measurement of whole-body extracellular volume (ECV) is of considerable clinical interest.

Conventional assessments of volume status, including physical examination, body weight, and central venous pressure, are imprecise and poorly correlated with true intravascular or interstitial fluid volumes. Indicator-dilution techniques using ions or isotopes^5-7^, as well as kinetic analysis of intravenously administered filtration markers like ^51^Cr-labeled ethylenediaminetetraacetic acid^4,8^, inulin analogs^9^ or the radiographic contrast agent iohexol^10-12^ have long served as gold standards for direct ECV measurement. However, their clinical use is limited by methodological complexity and reliance on somewhat cumbersome plasma sampling. By comparison, while bioimpedance is relatively simple, inexpensive and accessible, it has limited accuracy and precision^13^.

Importantly, the distribution of such indicators across extracellular compartments occurs at different rates. The combined plasma and interstitial space is herein referred to as the fast-exchange (FE) ECV. Equilibration into the FE-ECV typically occurs within seconds to minutes. In contrast, indicator distribution into the more slowly equilibrating “gel phase” or gel-phase compartment, herein referred to as the slow-exchange (SE) ECV, occurs over a longer time course^14-17^. This compartment is sometimes referred to as the “third space”, although greatly varying definitions have been applied to the term third space.

Clinically used iodinated CT contrast agents rapidly distribute throughout the vascular and extravascular extracellular fluid compartments without penetrating into the intracellular space or binding to cellular membranes^18-20^. Advances in dual-energy CT and photon-counting CT (PCCT) have enabled quantitative imaging applications based on these properties, including CT-based quantification of glomerular filtration rate^21-23^ and tissue ECV assessment in the myocardium^24-26^, liver^27,28^ and tumors^29,30^. Whole-body ECV assessment using CT, however, has remained limited. As patients referred for TAVR routinely undergo pre-procedural CT imaging, we introduce a PCCT-based method for whole-body FE-ECV quantification. This study provides an initial clinical application, defining reference values in healthy subjects and comparing them with a TAVR cohort. We hypothesize that FE-ECV is increased in patients referred for TAVR compared with healthy reference values.

## Materials and Methods

### Study design and population

We conducted a prospective, single-center study involving two cohorts. The first cohort included healthy individuals scheduled for elective multiphase contrast-enhanced CT scans as part of the living kidney donor evaluation process between July 2022 and January 2024.

These participants were enrolled after providing written informed consent. The second cohort comprised patients enrolled in the Alpha Registry (NCT07016477) who referred for TAVR and underwent PCCT scanning for procedural planning between May 2024 and December 2025. Exclusion criteria included participants with metal implants in the abdominal region, absence of late-phase scans containing the whole urinary bladder with a scan delay between 6-10 minutes, lack of venous hematocrit measurement, and absence of late phase scan or spectral reconstruction (**Figure 1**). The study was approved by the Scientific and Research Ethics Committee of the Hungarian Medical Research Council (SE RKEB 18/2022 and BM/9095-1/2024) and was carried out in accordance with the Declaration of Helsinki.

**Figure 1.**
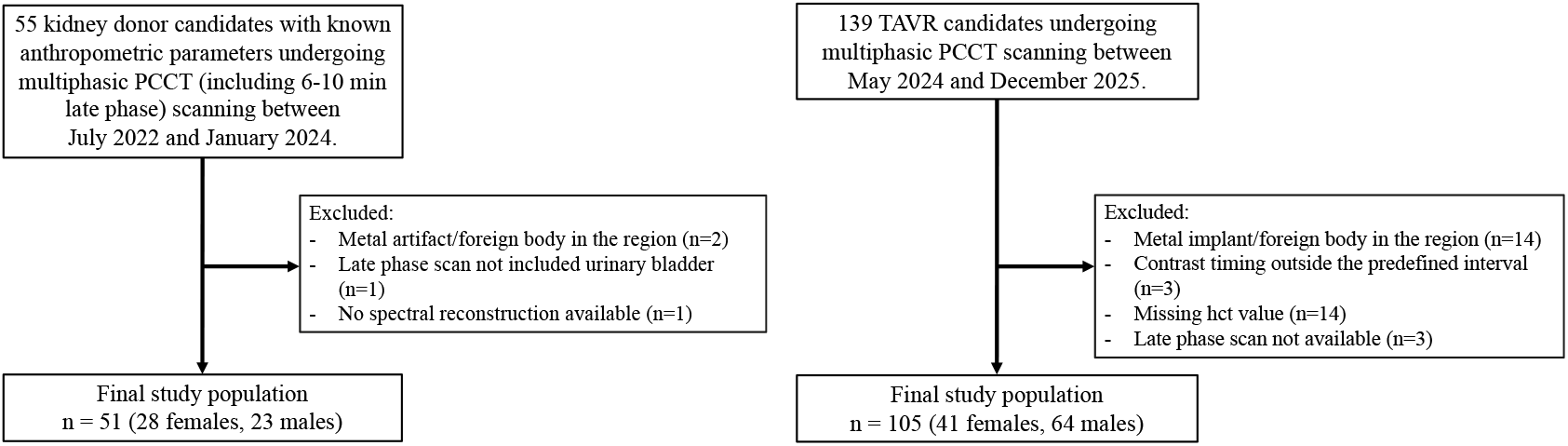
Study flowchart with inclusion and exclusion criteria. PCCT: Photon-counting computed tomography, TAVR: transcatheter aortic valve replacement.

### CT data acquisition and analysis

CT urography was performed using a dual-source PCCT (NAEOTOM Alpha.Peak, Siemens Healthineers, Forchheim, Germany). Iomeprol or iopromide was used as an intravenous contrast agent (Iomeron 350 or 400, Bracco Imaging Ltd, or Ultravist 370, Bayer Healthcare), and was administered as 42-120 mL volume at a flow rate of 3-5.5 mL/s from antecubital vein access via an 18-gauge catheter. Spectral CT data were acquired at 120 kVp (healthy controls) and 140 kVp (TAVR group). Helical acquisition with automated tube current modulation was performed with image quality levels of 100 and 145, respectively. Multiplanar reconstructions and iodine maps were created from late phase scans with a 2-mm slice thickness acquired at one time point 6-10 minutes after intravenous contrast administration, using commercially available software (Syngo.via, Siemens Healthineers, Forchheim, Germany). A 6–10 minute delay after contrast injection was selected to ensure adequate contrast equilibration within the FE-ECV compartment and to account for early renal excretion, while accommodating physiologic variability and workflow considerations. This interval also permitted evaluation of the impact of timing variations on FE-ECV quantification.

### Quantification of whole-body FE-ECV

Whole-body FE-ECV in mL was calculated using the following formula:

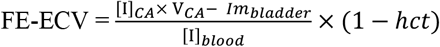

incorporating known injected iodinated contrast concentration ([I]_CA_, mg/mL) and volume (V_CA_, mL), measured blood iodine concentration ([I]_blood_, mg/mL), and urinary bladder total excreted iodine mass (Im_bladder_, mg) quantified on iodine maps from a region of interest in the inferior vena cava and regions of interest in all images covering the whole urinary bladder (**Figure 2**). For the measurement of bladder iodine mass, stacks covering the entire bladder with a slab thickness of 10 mm with no overlap in the sagittal plane were used. Segmentation was performed independently by two observers (one radiologist with 6 years of expertise and one radiologist in training with 4 years of expertise). Calculations included hematocrit measured from venous blood sampling. Boer’s formula was used to estimate lean body mass (LBM) based on height and weight.

**Figure 2.**
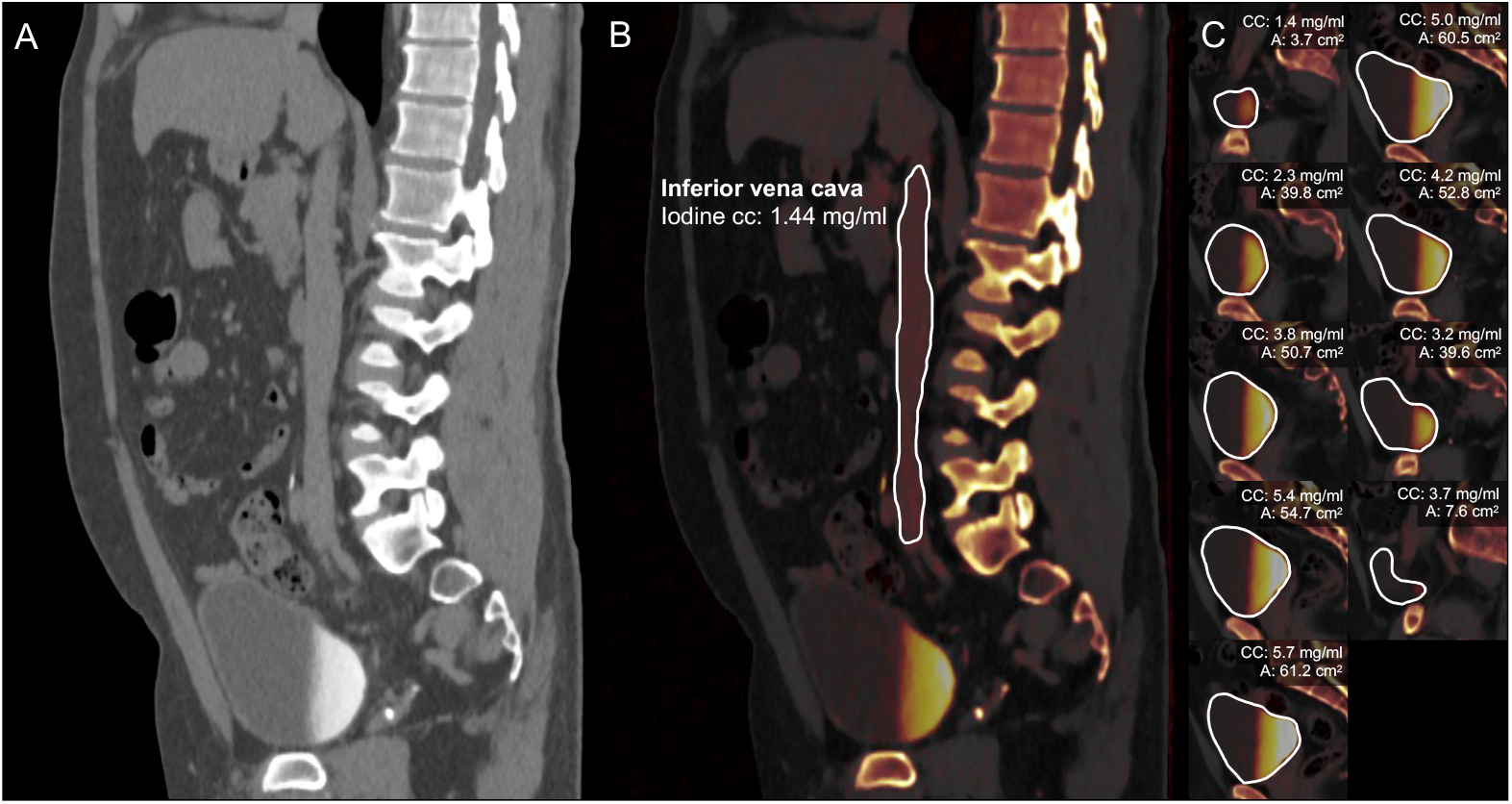
Photon-counting CT quantification of whole-body FE-ECV. Representative example of a late phase scan acquired with photon-counting CT (A) and the matching iodine map overlayed on virtual noncontrast series (B,C) with 10 mm slab thickness. This healthy male subject in his 30’s was scanned after an intravenous injection of 63 mL iomeprol (Iomeron 350), with a scan delay time of 7.9 minutes after contrast injection. The hematocrit was 46%, blood iodine concentration measured from the iodine map was 1.44 mg/mL (B), and total bladder iodine mass measured across all slices covering the bladder (C) was 1613 mg. Photon-counting CT quantified whole-body fast-exchange extracellular volume (FE-ECV) was 8.49 L and 130 mL/kg LBM. A: area, CC: concentration (iodine).

### Statistical analysis

Statistical analysis was conducted using R 4.1.1 (R Foundation for Statistical Computing, Vienna, Austria) and Stata 18.0 (StataCorp LLC College Station, TX). Continuous variables are reported as mean values ± standard deviation. Comparisons between continuous variables were performed using two-tailed Student’s t test while categorical variables were compared with Fisher’s exact test. The association between FE-ECV and patient or CT parameters was assessed using linear regression. Interobserver variability of iodine map measurements was evaluated in 20 randomly selected subjects using Bland-Altman analysis and the intraclass correlation coefficient (ICC). A two-tailed p-value <0.05 was considered statistically significant.

## Results

### Study population

Among healthy subjects, exclusions were due to metal implants in the region of interest (n=2), unavailable spectral image reconstruction for the late-phase scan (n=1), and incomplete urinary bladder coverage (n=1). In the TAVR group, exclusions were due to implants (n=14), contrast timing outside the predefined interval (n=3), missing hematocrit values (n=14), and unavailable late-phase scans (n=3). A total of 51 healthy subjects and 105 patients referred for TAVR were included in the final analysis. Compared to the healthy cohort, TAVR patients were older (47±9 vs 78±6 years, p <0.001) and did not differ in proportion of female participants (55% vs 39%, p=0.09). Body mass index, body surface area, and estimated LBM was comparable between the two groups (all p>0.7). Participant characteristics are summarized in **Table 1**.

**Table 1.** Participant characteristics.

| **Patient characteristics**<br>Mean ±SD or n (%) | **Healthy subjects** | **TAVR patients** | **p** |
| --- | --- | --- | --- |
| Number of participants, n | 51 | 105 |  |
| Age, years | 47±9 | 78±6 | **<0.001** |
| Female sex, n (%) | 28 (55) | 41 (39) | 0.09 |
| Body weight, kg | 78±18 | 78±15 | 0.83 |
| Height, cm | 170±9 | 169±10 | 0.54 |
| Body mass index, kg/m<sup>2</sup> | 26.9±5.3 | 27.2±4.4 | 0.80 |
| Body surface area, m<sup>2</sup> | 1.89±0.23 | 1.88±0.21 | 0.74 |
| Estimated lean body mass, kg | 54.7±10.4 | 54.9±10.0 | 0.85 |
| Hematocrit, % | 42±3 | 35±6 | **<0.001** |
| eGFR, mL/min/1.73m<sup>2</sup> | 89±5 | 58±18 | **<0.001** |

### Normal values of PCCT-derived whole-body FE-ECV in healthy subjects

Table 2. shows CT-derived parameters contributing to FE-ECV calculation, stratified by cohort. FE-ECV was greater in healthy males compared to females (9.8±2.1 vs 7.9±1.6 L, p<0.001, **Figure 3**). However, FE-ECV did not differ between the sexes when indexed to LBM (154±21 vs 166±23 mL/kg LBM, p=0.07). Thus, the combined normal range was 160±22 mL/kg LBM, 95% limits 116–204 mL/kg LBM. FE-ECV correlated with LBM (R^2^=0.65, p<0.001, **Figure 3**), BMI (R^2^=0.41, p<0.001, not shown), and BSA (R^2^=0.69, p<0.001, not shown). However, when indexed to body weight, FE-ECV (mL/kg LBM) showed a small magnitude negative correlation with BMI (R^2^=0.18, p=0.02). FE-ECV indexed to LBM showed no correlation with eGFR or serum creatinine (p>0.12 for all). As shown on **Figure 4**. FE-ECV did not correlate with age (R^2^<0.01, p=0.81), or the time interval between the contrast injection and late phase imaging (R^2^<0.01, p=0.70), and did not differ between both types of injected contrast agent (p=0.66).

**Table 2.** CT measures of study subjects.

| **Measure**<br>Mean ±SD | **Healthy** | **TAVR** |
| --- | --- | --- |
| Injected contrast volume, mL | 66±15 | 103±15 |
| Injected iodine mass, mg | 23,323±5,213 | 40,491±6,469 |
| Injected iodine mass, mg/kg body weight | 300±26 | 541±142 |
| Time between contrast injection and late phase imaging, min | 8.2±0.8 | 7.5±1.5 |
| Blood iodine concentration, mg/mL | 1.45±0.21 | 2.74±0.61 |
| Excreted bladder iodine mass, mg | 1,473±545 | 1,702±776 |
| Excreted bladder iodine mass, % of injected | 6.3±1.9 | 4.2±1.8 |
| Late phase scan DLP (mGy x cm)* | 242.9±82.7 | 248.3±82.6 |

DLP: dose length product
\*The healthy cohort underwent a late-phase abdomen–pelvis scan, whereas the TAVR group underwent a pelvis-only scan, with additional abdominal coverage performed in selected cases when clinically indicated.

**Figure 3.**
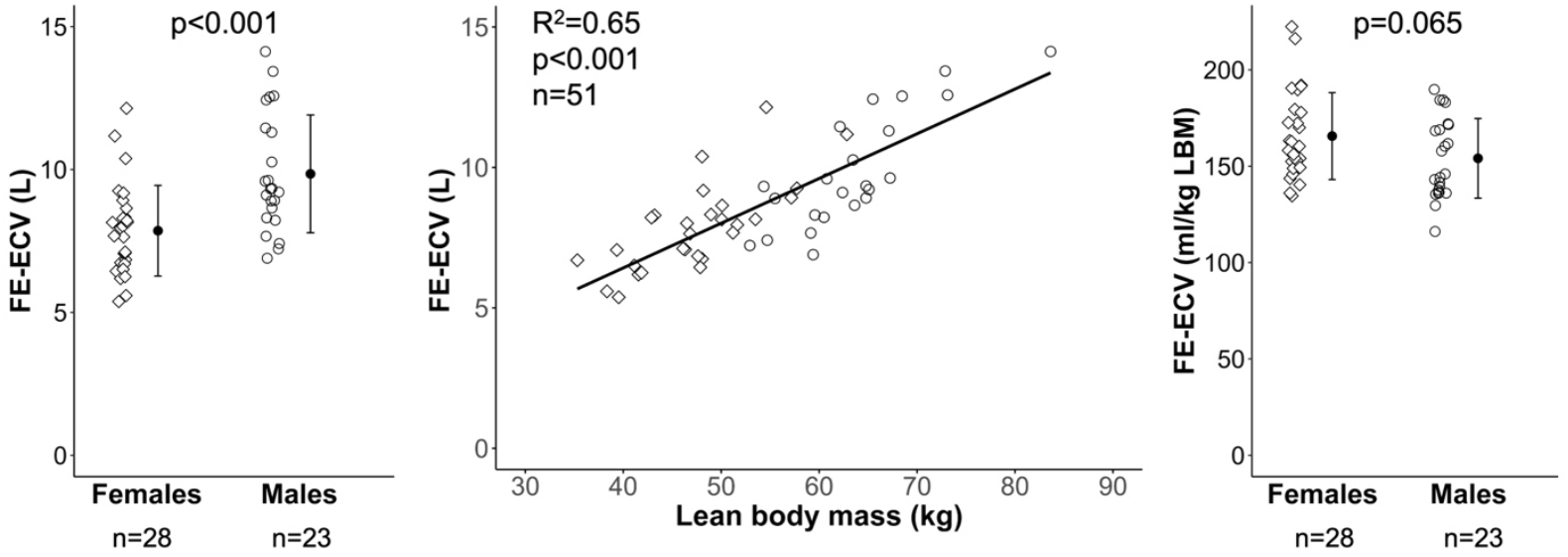
Whole-body fast-exchange extracellular volume (FE-ECV) quantified by photon-counting CT in healthy female and male subjects. Left: FE-ECV (L) is greater in males compared to females, and shows a strong correlation with lean body mass (Middle). Right: FE-ECV does not differ between the sexes when indexed to lean body mass.

**Figure 4.**
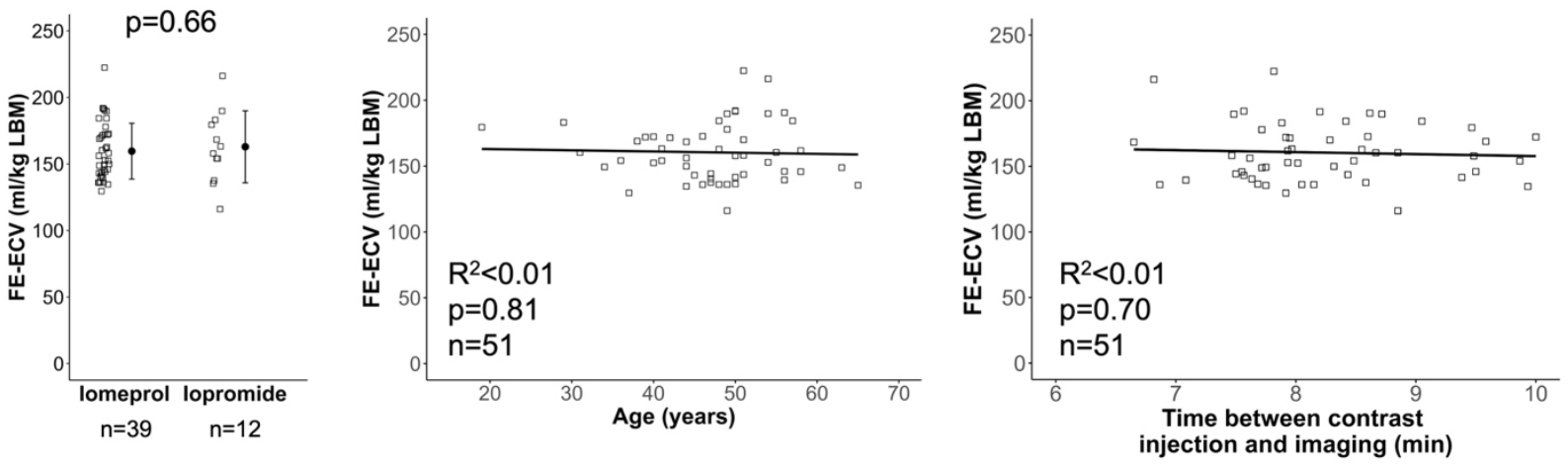
Whole-body FE-ECV is unaffected by contrast protocol or age in healthy controls. PCCT-derived whole-body fast-exchange extracellular volume (FE-ECV) is independent of (left) the type of injected iodinated contrast agent, (middle) subject age, and (right) the time interval between contrast administration and late-phase imaging. FE-ECV: fast-exchange extracellular volume, LBM: lean body mass.

### Interobserver variability of PCCT-measured ECV

The ICC [95%CI] of blood iodine concentration, bladder iodine mass and FE-ECV calculated from iodine maps were 0.96 [0.91-0.98], 1.00 [1.00-1.00], and 0.98 [0.94-0.99], respectively, indicating a good agreement between the two observers. Bland-Altman analysis showed a bias of 0.18±0.36 L in FE-ECV, -0.04±0.07 mg/mL in blood iodine concentration and 16.9±28.7 mg in bladder iodine mass between the two observers.

### Elevated PCCT-derived FE-ECV in TAVR candidates

Patients referred for TAVR demonstrated a higher mean whole-body FE-ECV compared with healthy subjects (9.4±1.8 vs 8.8±2.1 L, p=0.0495, and 174±34 vs 160 ± 22 mL/kg LBM, p=0.01; **Figure 5**.). FE-ECV in the group did not correlate with eGFR (R^2^=0.03, p=0.07).

**Figure 5.**
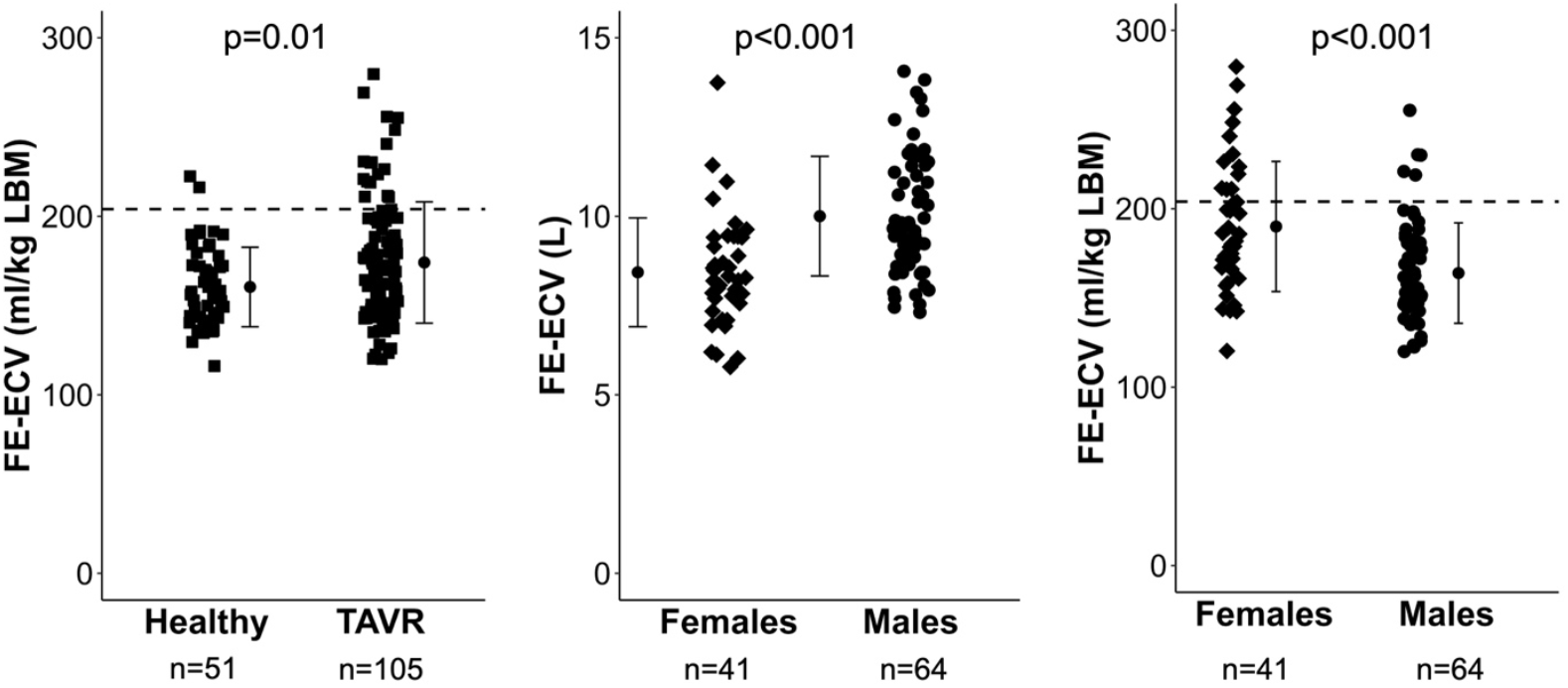
Markedly elevated FE-ECV in TAVR candidates with pronounced sex-specific differences. Fast-exchange extracellular volume (FE-ECV) is significantly increased in patients undergoing transcatheter aortic valve replacement (TAVR) compared with healthy controls (left). Absolute FE-ECV (L) is higher in males than in females (middle). However, when indexed to lean body mass (LBM), females demonstrate a higher FE-ECV (right). Dash line represents the upper limit of the healthy normal range (204 ml/kgLBM). FE-ECV: fast-exchange extracellular volume, LBM: lean body mass, TAVR: transcatheter aortic valve replacement.

Male patients had higher absolute FE-ECV (L) than female patients (10.0±1.7 vs 8.4±1.5 L, p<0.001), whereas indexing to LBM revealed higher FE-ECV in females (190±36 vs 164±28 ml/kg LBM, p<0.001). Overall, 19 patient (18%) of TAVR candidates exceeded the upper limit of the normal FE-ECV range (204 ml/kg LBM). Female patients were more likely to exceed this limit than male patients (21% vs 6%, p=0.007). Similar results were observed when using sex-specific cutoffs, with 20 patients exceeding the respective upper limits.

## Discussion

Patients with severe aortic stenosis are at increased risk of having fluid volume overload. In individuals referred for TAVR, excess extracellular fluid volume has been linked to worse clinical outcomes and its quantification may improve risk prediction beyond conventional stratification tools. In this study, we introduced a PCCT-based approach to quantify whole-body fast-exchange extracellular volume (FE-ECV) and evaluated FE-ECV status in healthy volunteers and patients undergoing TAVR.

This study provides four main findings. First, single timepoint, late-phase PCCT iodine mapping is an effective method for quantifying whole-body FE-ECV, unaffected by contrast type or dose, and scan delay, highlighting the robustness of the method. Second, sex-independent normal reference values of FE-ECV indexed to LBM are presented. Third, candidates for TAVR presented elevated PCCT-derived FE-ECV values, compared to healthy individuals. Finally, the elevated FE-ECV was more prevalent among female TAVR candidates in the study cohort, with higher lean body mass-indexed FE-ECV values, and a greater proportion exceeding the normal range compared with male patients, suggesting potential sex-specific differences in physiology, volume status and treatment response.

Almost one fifth of the TAVR candidates in this study exhibited elevated FE-ECV, with the average values also exceeding that of healthy controls. This prevalence is slightly lower than that reported in a prior study using bioimpedance spectroscopy, which indentified fluid overload in 40% of patients before TAVR, although only 28% of those subjects demonstrated pulmonary congestion on chest radiography^3^. Bioelectrical impedance spectroscopy is a relatively accessible, low-cost, non-invasive method. However, it has been shown to overestimate ECV compared to indicator dilution techniques^31,32^. Others have reported a fluid overload prevalence of 25% among TAVR candidates without prior cardiac decompensation, defined by the presence of pulmonary rales, peripheral edema, or pleural effusion. This prevalence increased to more than 50% in patients with a history of cardiac decompensation and reached 90% among those presenting with acute decompensation^33^. Taken together with our findings, these data suggest a substantial presence of subclinical decompensation among patients referred for TAVR.

Interestingly, 21% of the female TAVR candidates had a FE-ECV exceeding the normal range, while only 6% male patients had a FE-ECV above the threshold. Of note, although females generally derive similar clinical benefit from evidence-based heart failure drug and device therapies as men, the utilization rates of guideline-directed medical therapies remain lower in females^34^. In contrast, among healthy controls in this study, FE-ECV indexed to LBM did not differ between sexes, although a trend toward higher values was observed in females, consistent with prior reports using blood sampling-based indicator-dilution methods showing higher LBM indexed ECV values in females^35^. FE-ECV indexed to LBM was also independent of age, in line with previous reports^35,36^. In accordance with the literature, unindexed FE-ECV values were higher among males in both healthy controls and TAVR candidates^37^, and correlated with body size^36^.

Healthy PCCT-derived FE-ECV normal values presented in this study are approximately 40% lower than total ECV quantified using other indicator-dilution methods based on venous blood sampling^4,9,16,35,36,38,39^. In the current study, measurements were acquired 6-10 minutes after contrast injection, and the study aimed to quantify the FE-ECV compartment that is more dynamic and prone to volume changes, likely encompassing plasma volume and interstitial fluids resembling a plasma ultrafiltrate, into which ions and small molecules diffuse freely^16^. This approach excludes the SE-ECV compartments such as cerebrospinal fluid, serous, and synovial fluids, bile ducts, and gastrointestinal fluids. Furthermore, connective tissue fluid is also a “non-functional” fluid space with little or at least delayed diffusion equilibrium with the rest of the extracellular space, thus also comprising the SE-ECV^14,15^. These physiological differences across extracellular fluid spaces also explain variability in ECV estimates obtained with different tracers, such as the difference between the smaller “inulin space” measured by inulin or thiosulfate, and the usually larger “chloride space” measured by chloride, bromide, or ^51^Cr-EDTA^16^. It is likely that FE-ECV is more relevant to the clinical assessment of volume overload since not every compartment of the anatomical extracellular fluid spaces could harbour fluid overload-induced ECV changes. Within the limited sampling time window (6-10 minutes), FE-ECV quantification in our study was not dependent on precise scan timing or contrast agent type (iomeprol vs iomepromid). Importantly, the presented normal reference values provided a useful benchmark for identifying patients with volume expansion exceeding the normal range.

A limitation of our study is its single-center nature. However, consecutive patients were enrolled. The effects of different CT acquisition parameters, such as tube voltage were not tested in our present analysis. Due to the different scan indication and patient population, different contrast doses were used in the healthy participants and patients, respectively. However, healthy volunteers showed no difference in FE-ECV indexed to LBM over a range of contrast doses, suggesting that contrast dose was not an issue. FE-ECV derived from iodine mass balance is influenced by contrast kinetics and is not dependent on renal function, as reflected by the lack of correlation with eGFR. Furthermore, for healthy participants and the minority of TAVR candidates, venous sampling for hematocrit measurement and CT urography were not performed simultaneously. As this primarily affects healthy participants, the potential impact on the results is likely negligible. Another limitation of this study is the lack of gold-standard validation. Our investigation was designed as an initial clinical application of CT-based FE-ECV and did not assess its predictive value for clinical outcomes or compare it with standard TAVR risk stratification. Future studies should explore whether FE-ECV provides incremental prognostic information beyond simpler measures and examine its relationship with clinical parameters and biomarkers, including NT-proBNP, diuretic therapy, heart failure status, and echocardiographic findings.

In conclusion, the study has demonstrated that one in five patients referred for TAVR presented with increased FE-ECV, revealing a substantial prevalence of fluid overload quantified by single–time point late-phase PCCT iodine mapping. Excess FE-ECV was more frequently observed among female patients, suggesting that future studies should further investigate potential sex-specific differences in treatment and/or physiological differences in volume status and therapeutic response. Future longitudinal studies are needed to assess associations of CT-derived FE-ECV and adverse clinical outcomes. Overall, the PCCT-based approach presented in this study shows promise as a quantitative tool and biomarker for future applications in diagnosing and monitoring volume overload, and identifying patients with volume expansion.

## Data Availability

All data produced in the present study are available upon reasonable request to the authors

## Abbreviations

BMI: body mass index
FE-ECV: fast-exchange extracellular fluid volume
LBM: lean body mass
PCCT: photon-counting computed tomography
ROI: region of interest
TAVR: transcatheter aortic valve replacement

## References

1. Nitsche C, Kammerlander AA, Koschutnik M, et al. Fluid overload in patients undergoing TAVR: what we can learn from the nephrologists. ESC Heart Fail. Apr 2021;8(2):1408–1416. doi:10.1002/ehf2.13226

2. Nitsche C, Kammerlander Andreas A, Koschutnik M, et al. Volume Status Impacts CMR–Extracellular Volume Measurements and Outcome in AS Undergoing TAVR. JACC: Cardiovasc Imaging. 2021/02/01 2021;14(2):516–518. doi:10.1016/j.jcmg.2020.08.010

3. Halavina K, Koschutnik M, Donà C, et al. Quantitative fluid overload in severe aortic stenosis refines cardiac damage and associates with worse outcomes. Eur J Heart Fail. Oct 2023;25(10):1808–1818. doi:10.1002/ejhf.2969

4. Anand IS, Ferrari R, Kalra GS, Wahi PL, Poole-Wilson PA, Harris PC. Edema of cardiac origin. Studies of body water and sodium, renal function, hemodynamic indexes, and plasma hormones in untreated congestive cardiac failure. Circulation. 1989;80(2):299–305. doi:doi:10.1161/01.CIR.80.2.299

5. Ellis KJ, Wong WW. Human hydrometry: comparison of multifrequency bioelectrical impedance with2H2O and bromine dilution. J Appl Physiol. 1998;85(3):1056–1062. doi:10.1152/jappl.1998.85.3.1056

6. Tarazi RC, Dustan HP, Frohlich ED. Long-Term Thiazide Therapy in Essential Hypertension. Circulation. 1970;41(4):709–717. doi:10.1161/01.CIR.41.4.709

7. Binder C, Leth A. The distribution volume of 82Br-as a measurement of the extracellular fluid volume in normal persons. Scand J Clin Lab Invest. May 1970;25(3):291–7. doi:10.3109/00365517009046208

8. Faucon A-L, Flamant M, Metzger M, et al. Extracellular fluid volume is associated with incident end-stage kidney disease and mortality in patients with chronic kidney disease. Kidney Int. 2019/10/01/ 2019;96(4):1020–1029. doi:10.1016/j.kint.2019.06.017

9. Galatius S, Bent-Hansen L, Wroblewski H, Kastrup J. Plasma clearance of polyfructosan and extracellular body fluid distribution in idiopathic dilated cardiomyopathy and after heart transplantation. Am J Cardiol. Apr 1 2000;85(7):843–8. doi:10.1016/s0002-9149(99)00878-4

10. Bird NJ, Peters C, Michell AR, Peters AM. Extracellular distribution volumes of hydrophilic solutes used to measure the glomerular filtration rate: comparison between chromium-51-EDTA and iohexol. Physiol Meas. 2007/01/19 2007;28(2):223. doi:10.1088/0967-3334/28/2/010

11. Zdolsek JH, Lisander B, Hahn RG. Measuring the size of the extracellular fluid space using bromide, iohexol, and sodium dilution. Anesth Analg. 2005;101(6):1770–1777. doi:10.1213/01.Ane.0000184043.91673.7e

12. Bird NJ, Peters C, Michell AR, Peters AM. Association between glomerular filtration rate and extracellular fluid volume in normal subjects and patients with renal impairment. Scand J Clin Lab Invest. 2008/01/01 2008;68(1):39–49. doi:10.1080/00365510701444629

13. Schotman J, Rolleman N, van Borren M, et al. Accuracy of Bioimpedance Spectroscopy in the Detection of Hydration Changes in Patients on Hemodialysis. J Ren Nutr. 2023/01/01/ 2023;33(1):193–200. doi:10.1053/j.jrn.2021.11.004

14. Roumelioti ME, Glew RH, Khitan ZJ, et al. Fluid balance concepts in medicine: Principles and practice. World J Nephrol. Jan 6 2018;7(1):1–28. doi:10.5527/wjn.v7.i1.1

15. Walser M, Seldin DW, Grollman A. An evaluation of radiosulfate for the determination of the volume of extracellular fluid in man and dogs. J Clin Invest. Apr 1953;32(4):299–311. doi:10.1172/jci102739

16. Ljunggren H, Ikkos D, Luft R. Studies on body composition. I. Body fluid compartments and exchangeable potassium in normal males and females. Acta Endocrinol (Copenh). Jun 1957;25(2):187–198.

17. Cousins C, Gunasekera RD, Mubashar M, et al. Comparative kinetics of microvascular inulin and 99mTc-labelled diethylenetriaminepenta-acetic acid exchange. Clin Sci (Lond). Nov 1997;93(5):471–7. doi:10.1042/cs0930471

18. Lorusso V, Luzzani F, Bertani F, Tirone P, de Haën C. Pharmacokinetics and tissue distribution of iomeprol in animals. Eur J Radiol. May 1994;18 Suppl 1:S13–20. doi:10.1016/0720-048x(94)90090-6

19. Lorusso V, Taroni P, Alvino S, Spinazzi A. Pharmacokinetics and safety of iomeprol in healthy volunteers and in patients with renal impairment or end-stage renal disease requiring hemodialysis. Invest Radiol. Jun 2001;36(6):309–16. doi:10.1097/00004424-200106000-00002

20. Krause W, Schuhmann-Giampieri G, Staks T, Kaufmann J. Dose proportionality of iopromide pharmacokinetics and tolerability after IV injection in healthy volunteers. Eur J Clin Pharmacol. 1994/07/01 1994;46(4):339–343. doi:10.1007/BF00194402

21. Stehlé T, Wei F, Brabant S, et al. Glomerular Filtration Rate Measured Based on Iomeprol Clearance Assessed at CT Urography in Living Kidney Donor Candidates. Radiology. 2023;309(3):e230567. doi:10.1148/radiol.230567

22. Wang T, Xu Y, Liu W, et al. Measurement of Glomerular Filtration Rate Using Multiphasic Computed Tomography in Patients With Unilateral Renal Tumors: A Feasibility Study. Front Physiol. 2019;10:1209. doi:10.3389/fphys.2019.01209

23. Yuan X, Zhang J, Tang K, et al. Determination of Glomerular Filtration Rate with CT Measurement of Renal Clearance of Iodinated Contrast Material versus 99mTc-DTPA Dynamic Imaging “Gates” Method: A Validation Study in Asymmetrical Renal Disease. Radiology. 2017;282(2):552–560. doi:10.1148/radiol.2016160425

24. Nacif MS, Kawel N, Lee JJ, et al. Interstitial myocardial fibrosis assessed as extracellular volume fraction with low-radiation-dose cardiac CT. Radiology. Sep 2012;264(3):876–83. doi:10.1148/radiol.12112458

25. Cundari G, Galea N, Mergen V, Alkadhi H, Eberhard M. Myocardial extracellular volume quantification with computed tomography—current status and future outlook. Insights Imaging. 2023/09/25 2023;14(1):156. doi:10.1186/s13244-023-01506-6

26. Mergen V, Sartoretti T, Klotz E, et al. Extracellular Volume Quantification With Cardiac Late Enhancement Scanning Using Dual-Source Photon-Counting Detector CT. Invest Radiol. Jun 1 2022;57(6):406–411. doi:10.1097/rli.0000000000000851

27. Mizuno M, Tago K, Okada M, et al. Extracellular volume by dual-energy CT, hepatic reserve capacity scoring, CT volumetry, and transient elastography for estimating liver fibrosis. Sci Rep. Dec 12 2023;13(1):22038. doi:10.1038/s41598-023-49362-0

28. Tago K, Tsukada J, Sudo N, et al. Comparison between CT volumetry and extracellular volume fraction using liver dynamic CT for the predictive ability of liver fibrosis in patients with hepatocellular carcinoma. Eur Radiol. Nov 2022;32(11):7555–7565. doi:10.1007/s00330-022-08852-x

29. Wang X, Du L, Cao Y, et al. Comparing extracellular volume fraction with apparent diffusion coefficient for the characterization of breast tumors. Eur J Radiol. Feb 2024;171:111268. doi:10.1016/j.ejrad.2023.111268

30. Fukukura Y, Kumagae Y, Higashi R, et al. Extracellular volume fraction determined by equilibrium contrast-enhanced dual-energy CT as a prognostic factor in patients with stage IV pancreatic ductal adenocarcinoma. Eur Radiol. Mar 2020;30(3):1679–1689. doi:10.1007/s00330-019-06517-w

31. Armstrong LE, Kenefick RW, Castellani JW, et al. Bioimpedance spectroscopy technique: intra-, extracellular, and total body water. Med Sci Sports Exerc. Dec 1997;29(12):1657–63. doi:10.1097/00005768-199712000-00017

32. Simpson JAD, Lobo DN, Anderson JA, et al. Body water compartment measurements: A comparison of bioelectrical impedance analysis with tritium and sodium bromide dilution techniques. Clin Nutr. 2001;20(4):339–343. doi:10.1054/clnu.2001.0398

33. Fischer-Rasokat U, Renker M, Charitos EI, et al. Cardiac decompensation of patients before transcatheter aortic valve implantation—clinical presentation, responsiveness to associated medication, and prognosis. Original Research. Front Cardiovasc Med. 2023-October-23 2023;Volume 10-2023 doi:10.3389/fcvm.2023.1232054

34. Crousillat DR, Ibrahim NE. Sex Differences in the Management of Advanced Heart Failure. Curr Treat Options Cardiovasc Med. 2018/09/21 2018;20(11):88. doi:10.1007/s11936-018-0687-y

35. Peters AM, Perry L, Hooker CA, et al. Extracellular fluid volume and glomerular filtration rate in 1878 healthy potential renal transplant donors: effects of age, gender, obesity and scaling. Nephrol Dial Transplant. Apr 2012;27(4):1429–37. doi:10.1093/ndt/gfr479

36. Brøchner-Mortensen J. The extracellular fluid volume in normal man determined as the distribution volume of [51Cr] EDTA. Scand J Clin Lab Invest. 1982/01/01 1982;42(3):261–264. doi:10.1080/00365518209168083

37. Ritz P, Vol S, Berrut G, Tack I, Arnaud M, Tichet J. Influence of gender and body composition on hydration and body water spaces. Clin Nutr. 09/01 2008;27:740–6. doi:10.1016/j.clnu.2008.07.010

38. Faucon A-L, Flamant M, Delanaye P, et al. Estimating Extracellular Fluid Volume in Healthy Individuals: Evaluation of Existing Formulae and Development of a New Equation. Kidney Int Rep. 2022;7(4):810–822. doi:10.1016/j.ekir.2022.01.1057

39. Bird NJ, Peters AM. New gender-specific formulae for estimating extracellular fluid volume from height and weight in adults. Nucl Med Commun. Jan 2021;42(1):58–62. doi:10.1097/mnm.0000000000001301

